# Efficacy of Nirmatrelvir/ritonavir in reducing the risk of severe outcome in patients with SARS-CoV-2 infection: a real-life full-matched case-control study (SAVALO Study)

**DOI:** 10.1101/2024.06.07.24307681

**Authors:** I Gentile, A Giaccone, MM Scirocco, F Di Brizzi, F Cuccurullo, M Silvitelli, L Ametrano, FA Alfè, D Pietroluongo, I Irace, MR Chiariello, N De Felice, S Severino, G Viceconte, N Schiano Moriello, AE Maraolo, AR Buonomo, R Scotto, Federico II COVID team

**Author notes:** Federico II COVID Team is listed in the acknowledgement. **Corresponding author**: Dr. Agnese Giaccone. AORN Ospedali dei Colli, Cotugno Hospital, Department of Infectious Diseases, Unit of Geriatric Infectious Diseases, Via G. Quagliarello 54, 80131, Naples, Italy.

## Abstract

**Introduction:** Ritonavir-boosted nirmatrelvir (N/r) is an antiviral which targets the main viral protease, administered to prevent the progression of SARS-CoV-2 infection in patients at high risk for severe COVID-19. We present a real-life case-control study evaluating the efficacy of N/r therapy in SARS-CoV-2 omicron variants positive outpatients in Campania region, Italy, with the aim of assessing the occurrence of three outcomes (hospital admission, admission in ICU and death) in cases and controls.

**Methods:** We enrolled SARS-CoV-2 positive subjects that came to our attention in Early antiviral treatment ambulatory of Infectious Disease ward of University Federico II of Naples, Italy from January 1^st^, 2022, to December 31^st^, 2022, during the first five days from symptoms occurrence. Patients were enrolled as cases or controls if they were treated with N/r or not treated at all, respectively.

**Results:** 1064 patients were included (cases: 423, controls: 1184). Cases showed a lower mortality compared with controls while no differences were observed for other outcomes. Vaccinated patients showed a lower mortality compared with non-vaccinated ones (0.5% vs 7.8%, p<0.001). After full-matching propensity score, N/r reduced hospitalization rate only in unvaccinated patients. In contrast N/r significantly reduced mortality regardless of vaccination status.

**Conclusions:** Treatment with N/r has proven effective in reducing mortality among outpatients with SARS-CoV-2 infection during several omicron variant surges. The beneficial effect of N/r treatment in reducing progression is more pronounced in unvaccinated patients.

## Introduction

Coronavirus disease 2019 (COVID-19) is caused by the β-coronavirus SARS-CoV-2 which first emerged in Wuhan, China, in late 2019. It rapidly spread worldwide, resulting in over 774 million cases and 7 million deaths to date (1, 2). Early oral antiviral treatment is a crucial strategy in managing mild to moderate COVID-19, as it prevents the deterioration of patients, particularly those at high risk of developing severe disease (3). Nirmatrelvir is an orally administered antiviral agent that targets the SARS-CoV-2 3-chymotrypsin–like cysteine protease enzyme (Mpro), a key component of SARS-CoV-2 viral replication. In the EPIC-HR trial, treatment with ritonavir-boosted nirmatrelvir within 5 days of symptom onset resulted in 87.8% reduction in the risk of progression to severe disease compared to placebo. This was observed among non-hospitalized, unvaccinated adults at high risk of serious illness during the pre-Delta and Delta (B.1.617.2) pandemic waves (4).

However, the Omicron lineage variants and subvariants of SARS-CoV-2 have progressively displaced the previous variants due to their higher transmission rates and ability to evade the immune system. Despite this, N/r has maintained *in-vitro* activity, largely due to the low mutation rate of the Mpro gene (5). However gathering real-world data on its effectiveness against the emergent Omicron variants, which have been predominant since the beginning of 2022 (6), is now a research priority. It is also important to note that the population sample enrolled in the phase 3 trial was unvaccinated, contrasting with the current population, which is predominantly vaccinated or possesses hybrid immunity. This difference could potentially impact on the effectiveness of treatment in the real world. Aim of the present study is to assess the occurrence of hospital admission, admission in ICU and death in cases and controls, including patients with previous SARS-CoV-2 immunity, and to evaluate the effectiveness of N/r against the emergent Omicron variants.

## Methods

### Study design and population

This study was conducted as a retrospective case-control study. The eligibility criteria for participants were as follows: they had to be outpatients who tested positive for SARS-CoV-2 from January 2022 to December 2022, and they had to be aged 18 years or older. Subjects who were undergoing other therapies for COVID-19 (such as molnupiravir, remdesivir, sotrovimab, tixagevimab/cilgavimab or other anti-SARS-CoV-2 monoclonal antibodies), even in combination with N/r, were excluded from the study.

Cases in this study were defined as participants who sought care at the outpatient service of the Infectious Diseases Unit of the University Federico II of Naples, Italy, and received N/r within 5 days of symptoms onset. All treated patients had risk factors for disease progression as outlined by the Italian Drug Agency. These risk factors include being over 65 years of age, having a solid or haematological cancer, immunodeficiency, chronic liver or kidney disease, cardiovascular disease, uncontrolled diabetes mellitus, haemoglobinopathies, neurological disorders, chronic bronchopneumopathy, or severe obesity (BMI ≥30) (7).

Control participants were selected from a regional database that included all patients who tested positive for SARS-CoV-2 during the study period and did not receive any antiviral treatment. These control participants were identified and confirmed through telephone interviews conducted by healthcare professionals.

### Data collection and definitions

We collected information on patients’ demographic characteristics (age, sex), comorbid conditions (such as diabetes mellitus, hypertension, heart disease, lung disease, immunodeficiency, renal disease, neurological disease), vaccination status, and the predominant variant in our area during each segment of the study period. Patients were classified as fully vaccinated if they had received at least two doses of a COVID-19 vaccine.

For each patient, we calculated the Monoclonal Antibody Screening Score (MASS). This score was originally developed to quickly identify and stratify patients who should be prioritized for monoclonal antibody administration according to their risk of hospitalization (8). Additionally, we created a simplified Comorbidity Score. This score was assigned as follows: 2 points were given if the patient had at least 3 comorbidities, 1 point was given if the patient had 1 or 2 comorbidities, and 0 points were given if the patient had no comorbidity.

### Propensity score matching

We performed a propensity score (PS)–matched analysis to minimize selection bias and ensure an even distribution of confounding variables between the two groups under investigation: those treated with N/r and those untreated (receiving no treatment against COVID-19 at all) (9).

In conducting and reporting our PS methods, we adhered to the recommendations provided by Eikenboom et al. These guidelines aim to improve the quality of research on the effectiveness of antimicrobial therapy (10).

We estimated a PS model. Each subject was assigned a probability of receiving N/r based on his baseline characteristics. These characteristics included: age, sex, vaccination status, comorbid conditions (such as diabetes mellitus, hypertension, heart disease, lung disease, immunodeficiency, renal disease, neurological disease), MASS and Comorbidity score, and the predominant variant. Probabilities were estimated through probit regression. The balance of PS matching was evaluated using the absolute standardized mean difference (SMD) of covariates (both continuous and categorical) among groups. We considered a value less than 0.1 to represent acceptable balance. We assessed several matching algorithms, including greedy (with different ratios and calipers), optimal, and full (11). The best balance, retaining the entire recruited sample size, was achieved with full matching. This method assigned all units in the sample to one subclass each, either containing one treated unit and one or more control units, or one control unit and one or more treated units. The chosen number of subclasses and the assignment of units to subclasses minimized the sum of the absolute within-subclass distances in the matched sample (12).

### Outcomes

The primary outcomes of the study were as follows:

- The proportions of hospital admission, intensive care unit (ICU) admission, and 28-day all-cause mortality among both the treated individuals and the propensity-matched untreated individuals.
- A composite outcome was also evaluated, which was the occurrence of at least one of the following: hospital admission, ICU admission and all-cause death.

The follow-up duration was 28 days since symptom onset or first positive test for SARS-CoV-2.

### Statistical analysis

In this study, categorical variables are presented as numbers and percentages, while continuous variables are presented as median values and interquartile ranges (IQRs). The Mann-Whitney U test was used to determine differences in baseline characteristics between groups for continuous variables, and Pearson’s χ2 test was used for categorical variables.

After matching, the effect of N/r on the outcomes under investigation was estimated. The chosen estimand was the average treatment effect on the treated (ATT), which represents the average effect of treatment for those who receive treatment. This was done to answer the following archetypical question: should medical providers withhold treatment from those currently receiving it (13)?

Marginal effects, which are the comparisons between the expected potential outcome under treatment and the expected potential outcome under control, were estimated and were expressed as odds ratios (ORs) with their 95% confidence intervals (CIs). Estimation was performed through g-computation with a cluster-robust standard to account for pair membership, taking into account binary outcomes with covariates by means of a logistic regression (14). Including covariates in the outcome model after matching serves several functions: it can increase precision in the effect estimate, reducente the bias due to residual imbalance, and make the effect estimate “doubly robust”. This means the estimate is consistent if either the matching reduces sufficient imbalance in the covariates or if the outcome model is correct (15).

Analyses were performed using R, version 4.1.0 (R Core Team), with the following packages:

#### MatchIt, cobalt, marginal effects

### Secondary analysis

We conducted a moderation analysis to determine if the treatment effect varied across different levels of another variable, specifically the vaccination status. The aim was to achieve balance within each subgroup of the moderating variable. This was done by performing matching in the full dataset (16).

### Ethical Review

This study was approved by the ethics committee of our academic hospital (protocol number 0015191, 22nd March 2023). The informed consent was collected for all patients included in the study. The informed consent from control patients was obtained via telephone interview. This method of consent collection was thoroughly reviewed by the ethics committee, ensuring that it met all ethical standards. The informed consent from case patients was written. As a result, all telephone conversations, including the verbal consent given by the patients, were recorded and securely stored. These records can be retrieved and reviewed if requested by any relevant authority.

## Results

The study included a total of 1,607 patients who met the inclusion/exclusion criteria. This comprised 423 cases (26.3%) and 1184 controls (73.7%). The baseline characteristics of these cases and controls are detailed in *Table 1*. Patients treated with N/r had a more complex clinical profile. Specifically, they were older (p<0.001), had obesity more frequently (p<0.001) and more often had immunodeficiency (p<0.001). These patients also exhibited a higher comorbidity score (p<0.001) and a higher MASS score (p<0.001). Conversely, patients in the control group had a higher proportion of chronic heart disease (p<0.001).

**Table 1.** Baseline features of pre-matched sample (N=1607).

|  | **Nirmatrelvir/ritonavir** | **No therapy** | **p-value** |
| --- | --- | --- | --- |
| N | 423 | 1184 |  |
| Male Sex (n, %) | 202 (47.8) | 524 (44.3) | 0.237 |
| Age (median [IQR]) | 63.00 [50.50, 72.50] | 57.00 [46.00, 67.00] | <0.001 |
| Vaccination (%) | 408 (96.5) | 1096 (92.6) | 0.007 |
| Diabetes (%) | 39 (9.2) | 102 (8.6) | 0.781 |
| Hypertension (%) | 12 (10.8) | 243 (20.5) | 0.020 |
| Chronic heart disease (%) | 35 (8.3) | 196 (16.6) | <0.001 |
| COPD (%) | 33 (7.8) | 75 (6.3) | 0.357 |
| CKD (%) | 8 (1.9) | 23 (1.9) | 1.000 |
| Obesity (%) | 85 (20.1) | 130 (11.0) | <0.001 |
| Liver disease (%) | 2 (0.5) | 11 (0.9) | 0.560 |
| Neurological disease (%) | 14 (3.3) | 44 (3.7) | 0.816 |
| Immunodeficiency (%) | 209 (49.4) | 63 (5.3) | <0.001 |
| Comorbidity Score (median [IQR]) | 1.00 [1.00, 1.00] | 0.00 [0.00, 1.00] | <0.001 |
| MASS score (median [IQR]) | 3.00 [2.00, 5.00] | 0.00 [0.00, 3.00] | <0.001 |
| Predominant variant (%) |  |  | <0.001 |
| - omicron (not specified) | 3 (0.7) | 116 (9.8) |  |
| - omicron_BA.1 | 45 (10.6) | 208 (17.6) |  |
| - omicron_BA.2 | 164 (38.8) | 260 (22.0) |  |
| - omicron_BA.5 | 201 (47.5) | 599 (50.6) |  |
| - omicron_BQ.1 | 5 (1.2) | 0 (0.0) |  |
| - omicron_XBB.1.5 | 5 (1.2) | 1 (0.1) |  |

*IQR: interquartile range; COPD: chronic obstructive pulmonary disease; CKD: chronic kidney disease*

In total, 27 patients (1.7%) required hospitalization and 2 (0.1%) were admitted to ICU. Remarkably, none of the 423 patients treated with N/r died. In contrast, 15 (1.3%) patients among controls died (p<0.05). However, there were no significant differences in the rates of hospital admission, ICU admission and in the composite outcome between the treated and untreated groups (Table 2)

**Table 2.**
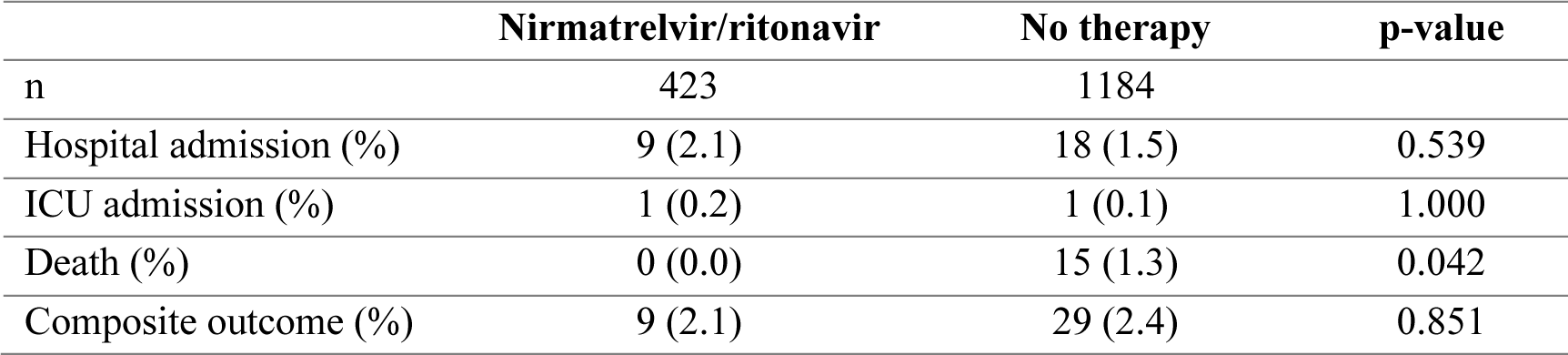
Outcome rates among cases and controls.

Following the full-matching PS analysis (see Supplementary material for covariate balance and distribution of PS between groups) it was confirmed that treatment with N/r significantly and drastically reduced the risk of death among patients with SARS-CoV-2 infection (*Table 3)*. Furthermore, N/r was found to significantly reduce the risk of the composite outcome. However, no effect of N/r on hospital admission alone was observed.

**Table 3.** Effect of N/r on different outcomes after full-matched propensity score.

|  | **Point estimate (OR)** | **Standard Error** | **95% Confidence Interval** | **P value** |
| --- | --- | --- | --- | --- |
| Composite outcome | 0.293 | 6.3 | 0.111-0.772 | 0.013 |
| Admission | 0.674 | 0.9 | 0.198-2.29 | 0.528 |
| ICU admission | * | * | * | * |
| Death | $2.6 \times 10^{-8}$ | 725.7 | $8.81 \times 10^{-9}$ - $7.69 \times 10^{-8}$ | <0.001 |

\*Algorithm did not converge (too few events).

**Table 4.** Outcome rates among included patients according to vaccination status.

|  | **Vaccination** | **No vaccination** | **p-value** |
| --- | --- | --- | --- |
| n | 1504 | 103 |  |
| Hospital admission (%) | 25 (1.7) | 2 (1.9) | 1.000 |
| ICU admission (%) | 2 (0.1) | 0 (0.0) | 1.000 |
| Death (%) | 7 (0.5) | 8 (7.8) | <0.001 |
| Composite outcome (%) | 30 (2.0) | 8 (7.8) | 0.001 |

When comparing outcomes based on vaccination status, it was found that patients who had received the SARS-CoV-2 vaccine had a significant lower rate of death (p<0.001). They also had a significantly lower rate of the composite outcome (p=0.001)

In the subgroup analysis conducted among both cases and control who received the SARS-CoV-2 vaccine, the effect of N/r on reducing the risk of death was confirmed in both vaccinated and unvaccinated subjects after full-matched propensity score analysis (*Table 5).* However, N/r was found to be effective in reducing the risk of hospital admission (p<0.001) and the risk of the composite outcome (p<0.001) only in non-vaccinated individuals.

**Table 5.** Effect of N/r on different outcomes after full-matched propensity score, according to vaccination status.

|  |  | Point estimate (OR) | Standard Error | 95% Confidence Interval | p-value | p subgroup |
| --- | --- | --- | --- | --- | --- | --- |
| Composite outcome | Vaccination Yes | 0.360 | 4.0 | 0.122 -1.060 | 0.0646 | <0.001 |
| | Vaccination No | $9.64 \times 10^{-7}$ | 337.7 | $2.72 \times 10^{-7}$ - $3.41 \times 10^{-6}$ | <0.001 | |
| Admission | Vaccination Yes | 0.578 | 1.1 | 0.138 - 2.42 | 0.453 | <0.001 |
| | Vaccination No | $1.29 \times 10^{-5}$ | 94.0 | $1.79 \times 10^{-6}$ - $9.26 \times 10^{-5}$ | <0.001 | |
| ICU admission | Vaccination Yes | * | * | * | * |  |
|  | Vaccination No | * | * | * | * |  |
| Death | Vaccination Yes | $5.27 \times 10^{-8}$ | 401.8 | $1.30 \times 10^{-8}$ - $2.14 \times 10^{-7}$ | <0.001 | 0.256 |
| | Vaccination No | $1.76 \times 10^{-8}$ | 557.6 | $4.98 \times 10^{-9}$ - $6.25 \times 10^{-8}$ | <0.001 | |

\*Algorithm did not converge (too few events).

## Discussion

In this retrospective matched cohort study, we evaluated the real-world effectiveness of N/r in reducing hospital admissions and mortality among patients with SARS-CoV-2 infection. We made comparisons by examining the outcomes of untreated individuals who were infected with SARS-CoV-2 during the same period. This was achieved using a full-matching propensity score, which helped ensure a fair and balanced comparison between the treated and untreated groups.

The majority of patients in this study contracted the infection during the surge of Omicron variants. Specifically, Omicron BA.2 and BA.5 were the most prevalent variants, with an estimated impact of 86,3% and 72.6% in cases and controls, respectively. Additionally, the vast majority of both cases and controls in our study (>90%) had received at least two doses of the SARS-CoV-2 vaccine. Given this high rate of vaccination and the prevalence of the Omicron subvariants, it is not surprising that the rates of hospitalization, ICU admission and mortality were remarkably low in line with worldwide epidemiological data (17–19). However, a significant finding was that all the 15 recorded deaths in this study occurred among the control group. This is a crucial result, particularly considering that the cases were considerably more vulnerable than the controls. Patients treated with N/r were indeed older than the controls and more frequently had obesity and immunodeficiency, resulting in higher comorbidity and MASS scores, as shown in Table 1. This outcome was confirmed after full matching PS analysis. The comparison analysis conducted after matching cases and controls also demonstrated a significant reduction in the composite outcome (at least one among hospital admission, ICU admission and death). Our results are consistent with those of other cohorts. For instance, Aggarwal et al. conducted a large propensity-matched, retrospective, observational cohort study of non-hospitalised patients infected with SARS-CoV-2 using records from a health system in Colorado, USA, between March and August 2022. This period corresponded to the widespread presence of the BA.4 and BA.5 variants. In this cohort N/r treatment was associated with a reduced 28-day all-cause hospitalisation compared with no antiviral treatment (adjusted odds ratio (OR) 0.45 [95% CI 0.33– 0.62]; p<0.0001) and reduced 28-day all-cause mortality (adjusted OR 0.15 [95% CI 0.03–0.50]; p=0.001) (5). Another study was conducted using health data records between November 2022 and March 2023 during which the predominant SARS-CoV-2 variants were BQ.1, BQ.1.1 and XBB.1.5. This study showed a similar efficacy of N/r treatment in preventing hospitalisation (20). In addition to the data from Aggarwal et al., our results also demonstrated a reduction in the risk of severe outcome (namely, the risk of the composite outcome: hospital admission, ICU admission, or death) among patients with SARS-CoV-2 treated with N/r.

Indeed, the value of real-world studies on N/r extends to evaluating its effectiveness among patients with prior immunity to SARS-CoV-2, whether acquired through vaccination or through natural infection. While the pivotal EPIC-HR trial only included unvaccinated patients, the landscape has significantly changed since then. Vaccination against COVID-19 has become widespread, with most of the general population having received at least two doses of the vaccine. As a result, some studies assessing the efficacy of nirmatrelvir-ritonavir have considered the impact of vaccination status on the outcomes in their multivariate analysis.

A real-world retrospective study published in September 2022 found that N/r demonstrated higher efficacy particularly among unvaccinated patients older than 65 years (21). Similar results were observed by Dryden-Peterson et al. in a cohort study conducted during the Omicron epidemic in a context of high vaccination prevalence. They reported an increased protective activity of N/r, with an 81% risk reduction among incompletely vaccinated individuals and patients who had received their most recent vaccine dose more than 20 weeks prior (19). In a population-based cohort study conducted in Quebec, Canada, a 96% reduction in the hospitalisation rate was reported among incompletely vaccinated outpatients (22). However, in this cohort, no benefit was observed among the vaccinated population, excluding severely immunocompromised patients and high-risk patients aged 70 years or older who had received their last vaccine dose more than 6 months prior. Despite this, further real-world studies have reported a maintained efficacy of N/r in preventing disease progression in subgroup analyses of patients who had received at least two doses of COVID-19 vaccine (23, 24).

In our cohort, the majority of patients (1,504, 93.6%) had received at least two doses of the SARS-CoV-2 vaccine. However, our analysis conducted after full-matching PS revealed that the effects of N/r were more pronounced on non-vaccinated patients. This was evidenced by a significant impact on hospital admission, death, and the composite outcome (see Table 5). However, it is important to emphasize that even among fully vaccinated patients, treatment with N/r significantly reduced mortality. This underscores the potential benefits of N/r treatment across different patient populations, regardless of their vaccination status.

The use of full-matching PS is a major strength of our study. Full matching can be viewed as a form of propensity score weighting that is less sensitive to the form of the propensity score model. This allows for the estimation of a weighted treatment effect that is ideally free of confounding by the measured covariates (25). One significant advantage of this matching algorithm is that all individuals are retained, often resulting in more balance than 1:1 matching (26). This was the case in our study, where there was a marked disproportion at baseline between the two groups in a key variable, namely the immunodeficiency status: 49.4% among treated and 5.3% among untreated, respectively. In contrast, the most commonly employed, nearest neighbor matching, would have resulted in a poor balance and also discarded a large number of observations, leading to a reduced power (27).

The central finding of our study is the positive impact of N/r on survival in a population where the majority (>90%) of patients were vaccinated. This contrasts with the recent findings form EPIC-SR study (28). This new randomized trial initially planned to exclude individuals with significant comorbidities, including immunosuppression, but later amended its protocol to include such participants if they had received a full course of vaccination. Notably, the EPIC-HR trial, which demonstrated a significant reduction in COVID-19–related hospitalization or death associated with N/r use compared to placebo, did not include vaccinated individuals (4). In the EPIC-SR study, where 56.9% of participants were vaccinated, no differences were detected between the N/r arm and placebo arm regarding time to sustained alleviation (the main outcome). However, in a planned subgroup analysis involving high-risk patients (all vaccinated with comorbidities) the relative reduction of N/r versus placebo in terms of hospitalization and death was 57% (3/317, 8/314), but this was not statistically significant (28). In contrast, our results showed a significant reduction in mortality among patients treated with N/r (Table 3). Furthermore, while we also observed the absence of efficacy of N/r in reducing the risk of hospitalization in the entire sample, we found that non-vaccinated patients treated with N/r showed a significant reduction in hospitalization risk compared with untreated subjects (Table 5). Even if a weak signal of benefit linked with N/r regarding major outcomes was detected in frail vaccinated subjects, for which the latest trial was not powered (29), a greater benefit can be inferred in unvaccinated high-risk patients. In our cohort, 10.7% of unvaccinated individuals had immunodeficiency (*Supplementary Table 1*). As is the case with many medical interventions, there is likely to be a gradient of benefit for the antiviral treatment, with the greatest benefit shown in the subjects at highest risk for progression (29).

We recognize that our study has several limitations. Firstly, our study is subject to all the biases inherent to its retrospective study design. While data for the cases were collected prospectively, data for the controls, who represent a significant percentage of all the included patients (73.7%), were retrieved retrospectively. Specifically, it is plausible that a proportion of the controls, identified through telephone interviews directly or via their relatives, may not have accurately reported baseline features (e.g., vaccination status) or outcomes. For instance, hospital admissions beyond the predefined follow-up window might have been reported as having occurred earlier due to recall bias. Despite efforts to track relatives of deceased patients, it is possible that a significant percentage of controls (or their relatives) who had an unfavourable outcome declined to be interviewed. However, given the large number of controls included in the study, this bias may have been mitigated. Moreover, we aimed to retrieve objective outcomes, discarding more subjective endpoints such as symptom resolution, which would have been difficult to reliably verify in the control group. It should also be noted that the collection of clinical data through telephone interviews, in the absence of reliable clinical files and interaction with healthcare professionals, compromises the trustworthiness and accuracy of such data. For this reason, the Charlson score was unsuitable for our analysis, and we had to identify a surrogate score to stratify each patient’s risk. Additionally, in our study, we were unable to trace back the time span since the last vaccine administration among the vaccinated population. Another limitation is the lack of detection of SARS-CoV-2 rebound, typically described as a recurrence of symptoms or a new positive viral test after testing negative, although definitions vary widely. Both interventional and observational studies have shown that subjects undergoing treatment with N/r may experience viral rebound (30). In fact, there is no consistent association between N/r and COVID-19 rebound, as it can occur after any other antiviral treatment and even in untreated subjects, likely due to a combination of factors such as immunosuppression, delayed viral clearance, and variable host-mounted immune response (31).

## Conclusions

N/r has been confirmed to effectively reduce the risk of death and severe outcomes in a population of patients primarily infected with the SARS-CoV-2 Omicron BA.2 and BA.5 variants. This population notably includes a significant proportion of immunosuppressed individuals. The efficacy of N/r is especially pronounced in non-vaccinated patients, where it has also been shown to decrease the risk of hospitalization. Interestingly, N/r has been found to significantly reduce mortality even in fully vaccinated patients. Given these findings, it is recommended that N/r should be promptly administered to all patients with early SARS-CoV-2 infection and risk factors for progression to severe COVID-19, irrespective of their vaccination status.

## Data Availability

The datasets generated and/or analysed during the current study, along with the Case Report Forms (CRFs), the written informed consent from case participants, and the records of the telephonic interviews conducted with control participants, including their verbal informed consent, are stored by the data manager at the Department of Clinical Medicine and Surgery, University of Naples Federico II, Via Sergio Pansini 5, 80131, Naples, Italy. These can be retrieved and reviewed upon request by any relevant authority.

## Acknowledgements

Federico II COVID Team: Francesco Antimo Alfè, Luigi Ametrano, Anna Borrelli, Antonio Riccardo Buonomo, Ferdinando Calabria, Giuseppe Castaldo, Letizia Cattaneo, Maria Rosaria Chiariello, Mariarosaria Cotugno, Federica Cuccurullo, Alessia d’Agostino, Noemi De Felice, Dario Diana, Francesco Di Brizzi, Giovanni Di Filippo, Isabella Di Filippo, Antonio Di Fusco, Federico Di Panni, Gaia Di Troia, Nunzia Esposito, Mariarosaria Faella, Lidia Festa, Maria Foggia, Maria Elisabetta Forte, Ludovica Fusco, Antonella Gallicchio, Gianpaolo Gargiulo, Ivan Gentile, Antonia Gesmundo, Agnese Giaccone, Carmela Iervolino, Irene Irace, Antonio Iuliano, Federica Licciardi, Giuseppe Longo, Matteo Lorito, Alberto Enrico Maraolo, Simona Mercinelli, Fulvio Minervini, Giuseppina Muto, Mariano Nobile, Daria Pietroluongo, Biagio Pinchera, Giuseppe Portella, Laura Reynaud, Alessia Sardanelli, Marina Sarno, Simone Severino, Maria Silvitelli, Nicola Schiano Moriello, Maria Michela Scirocco, Fabrizio Scordino, Riccardo Scotto, Stefano Mario Susini, Anastasia Tanzillo, Grazia Tosone, Emilia Trucillo, Ilaria Vecchietti, Giulio Viceconte, Emanuela Zappulo, Giulia Zumbo The final draft of this paper was revised with the assistance of Microsoft Copilot ©, powered by Chat-GPT 4.

## References

1. Hu B, Guo H, Zhou P, Shi ZL. Characteristics of SARS-CoV-2 and COVID-19. Nat Rev Microbiol. 2021;19(3):141–54.

2. WHO COVID-19 dashboard. Available at: https://data.who.int/dashboards/covid19/cases?n=c [last access April 2024].

3. Rinaldi M, Campoli C, Gallo M, Marzolla D, Zuppiroli A, Riccardi R, et al. Comparison between available early antiviral treatments in outpatients with SARS-CoV-2 infection: a real-life study. BMC infectious diseases. 2023;23(1):646.

4. Hammond J, Leister-Tebbe H, Gardner A, Abreu P, Bao W, Wisemandle W, et al. Oral Nirmatrelvir for High-Risk, Nonhospitalized Adults with Covid-19. The New England journal of medicine. 2022;386(15):1397–408.

5. Aggarwal NR, Molina KC, Beaty LE, Bennett TD, Carlson NE, Mayer DA, et al. Real-world use of nirmatrelvir-ritonavir in outpatients with COVID-19 during the era of omicron variants including BA.4 and BA.5 in Colorado, USA: a retrospective cohort study. The Lancet Infectious diseases. 2023;23(6):696-705.

6. GISAID hCoV-19 Variants Dashboard. Available at: https://gisaid.org/hcov-19-variants-dashboard/ [last access: March 2024].

7. AIFA RECOMMENDATIONS ON MEDICINES to be used in home management of COVID-19 cases. Vers. 7 – Updated 09/02/2022. Available at: https://www.aifa.gov.it/documents/20142/1269602/EN_Raccomandazioni_AIFA_gestione_domiciliare_COVID-19_Vers7_09.02.2022.pdf [last access: May 2024]

8. Razonable RR, Ganesh R, Bierle DM. Clinical Prioritization of Antispike Monoclonal Antibody Treatment of Mild to Moderate COVID-19. Mayo Clin Proc. 2022;97(1):26–30.

9. Thomas L, Li F, Pencina M. Using Propensity Score Methods to Create Target Populations in Observational Clinical Research. Jama. 2020;323(5):466–7.

10. Eikenboom AM, Le Cessie S, Waernbaum I, Groenwold RHH, de Boer MGJ. Quality of Conduct and Reporting of Propensity Score Methods in Studies Investigating the Effectiveness of Antimicrobial Therapy. Open Forum Infect Dis. 2022;9(4):ofac110.

11. Baek S, Park SH, Won E, Park YR, Kim HJ. Propensity score matching: a conceptual review for radiology researchers. Korean J Radiol. 2015;16(2):286–96.

12. Hansen BB, Klopfer SO. Optimal Full Matching and Related Designs via Network Flows. Journal of Computational and Graphical Statistics. 2006;15(3):609–27.

13. Li H, Wang C, Chen WC, Lu N, Song C, Tiwari R, et al. Estimands in observational studies: Some considerations beyond ICH E9 (R1). Pharm Stat. 2022;21(5):835–44.

14. Snowden JM, Rose S, Mortimer KM. Implementation of G-computation on a simulated data set: demonstration of a causal inference technique. American journal of epidemiology. 2011;173(7):731–8.

15. Nguyen TL, Collins GS, Spence J, Daurès JP, Devereaux PJ, Landais P, et al. Double-adjustment in propensity score matching analysis: choosing a threshold for considering residual imbalance. BMC Med Res Methodol. 2017;17(1):78.

16. Green KM, Stuart EA. Examining moderation analyses in propensity score methods: application to depression and substance use. J Consult Clin Psychol. 2014;82(5):773–83.

17. Hu FH, Jia YJ, Zhao DY, Fu XL, Zhang WQ, Tang W, et al. Clinical outcomes of the severe acute respiratory syndrome coronavirus 2 Omicron and Delta variant: systematic review and meta-analysis of 33 studies covering 6 037 144 coronavirus disease 2019-positive patients. Clinical microbiology and infection : the official publication of the European Society of Clinical Microbiology and Infectious Diseases. 2023;29(7):835–44.

18. Ward IL, Bermingham C, Ayoubkhani D, Gethings OJ, Pouwels KB, Yates T, et al. Risk of covid-19 related deaths for SARS-CoV-2 omicron (B.1.1.529) compared with delta (B.1.617.2): retrospective cohort study. Bmj. 2022;378:e070695.

19. Dryden-Peterson S, Kim A, Kim AY, Caniglia EC, Lennes IT, Patel R, et al. Nirmatrelvir Plus Ritonavir for Early COVID-19 in a Large U.S. Health System : A Population-Based Cohort Study. Annals of internal medicine. 2023;176(1):77–84.

20. Aggarwal NR, Beaty LE, Bennett TD, Fish LE, Jacobs JR, Mayer DA, et al. Real-world use of nirmatrelvir-ritonavir in COVID-19 outpatients during BQ.1, BQ.1.1., and XBB.1.5 predominant omicron variants in three U.S. health systems: a retrospective cohort study. Lancet Reg Health Am. 2024;31:100693.

21. Arbel R, Wolff Sagy Y, Hoshen M, Battat E, Lavie G, Sergienko R, et al. Nirmatrelvir Use and Severe Covid-19 Outcomes during the Omicron Surge. The New England journal of medicine. 2022;387(9):790–8.

22. Kaboré JL, Laffont B, Diop M, Tardif MR, Turgeon AF, Dumaresq J, et al. Real-World Effectiveness of Nirmatrelvir/Ritonavir on Coronavirus Disease 2019-Associated Hospitalization Prevention: A Population-based Cohort Study in the Province of Quebec, Canada. Clinical infectious diseases : an official publication of the Infectious Diseases Society of America. 2023;77(6):805–15.

23. Lewnard JA, McLaughlin JM, Malden D, Hong V, Puzniak L, Ackerson BK, et al. Effectiveness of nirmatrelvir-ritonavir in preventing hospital admissions and deaths in people with COVID-19: a cohort study in a large US health-care system. The Lancet Infectious diseases. 2023;23(7):806–15.

24. Low EV, Pathmanathan MD, Chidambaram SK, Kim WR, Lee WJ, Teh ZW, et al. Real-world nirmatrelvir-ritonavir outpatient treatment in reducing hospitalization for high-risk patients with COVID-19 during Omicron BA.4, BA.5 and XBB subvariants dominance in Malaysia: A retrospective cohort study. International journal of infectious diseases : IJID : official publication of the International Society for Infectious Diseases. 2023;135:77–83.

25. Austin PC, Stuart EA. The performance of inverse probability of treatment weighting and full matching on the propensity score in the presence of model misspecification when estimating the effect of treatment on survival outcomes. Statistical Methods in Medical Research. 2017;26(4):1654–70.

26. Austin PC. A comparison of 12 algorithms for matching on the propensity score. Stat Med. 2014;33(6):1057–69.

27. Stuart EA. Matching methods for causal inference: A review and a look forward. Stat Sci. 2010;25(1):1–21.

28. Hammond J, Fountaine RJ, Yunis C, Fleishaker D, Almas M, Bao W, et al. Nirmatrelvir for Vaccinated or Unvaccinated Adult Outpatients with Covid-19. The New England journal of medicine. 2024;390(13):1186–95.

29. Gandhi RT, Hirsch M. Treating Acute Covid-19 - Final Chapters Still Unwritten. The New England journal of medicine. 2024;390(13):1234–6.

30. Smith-Jeffcoat SE, Biddle JE, Talbot HK, Morrisey KG, Stockwell MS, Maldonado Y, et al. Symptoms, viral loads, and rebound among COVID-19 outpatients treated with nirmatrelvir/ritonavir compared to propensity score matched untreated individuals. Clinical infectious diseases : an official publication of the Infectious Diseases Society of America. 2023.

31. Petrosillo N. SARS-CoV-2 rebound with and without antivirals. The Lancet Infectious diseases. 2023;23(6):637–9.

