## Supplementary Table 1 for "Efficacy of Nirmatrelvir/ritonavir in reducing the risk of severe outcome in patients with SARS-CoV-2 infection: a real-life full-matched case-control study (SAVALO Study)"

**Supplementary Table 1.** Baseline features of pre-matched sample (N=1607) according to vaccination status.

| | **Vaccination** | **No vaccination** | **p-value** |
|---|---|---|---|
| N | 1504 | 103 | |
| Male sex (%) | 701 (46.6) | 25 (24.3) | <0.001 |
| Age (median, IQR) | 59.00 [47.00, 69.00] | 57.00 [44.50, 70.00] | 0.889 |
| Serology status (%) | | | 0.185 |
| - N/A | 1256 (83.5) | 93 (90.3) | |
| - positive | 163 (10.8) | 6 (5.8) | |
| - negative | 85 (5.7) | 4 (3.9) | |
| Diabetes (%) | 135 (9.0) | 6 (5.8) | 0.361 |
| Hypertension (%) | 248 (20.6) | 7 (7.8) | 0.005 |
| Chronic heart disease (%) | 222 (14.8) | 9 (8.7) | 0.123 |
| COPD (%) | 100 (6.6) | 8 (7.8) | 0.814 |
| CKD (%) | 31 (2.1) | 0 (0.0) | 0.271 |
| Obesity (%) | 207 (13.8) | 8 (7.8) | 0.114 |
| Liver disease (%) | 11 (0.7) | 2 (1.9) | 0.448 |
| Neurological disease (%) | 53 (3.5) | 5 (4.9) | 0.669 |
| Immunodeficiency (%) | 261 (17.4) | 11 (10.7) | 0.107 |
| Comorbidity score (median, IQR) | 1.00 [0.00, 1.00] | 0.00 [0.00, 1.00] | 0.001 |
| MASS score (median, IQR) | 2.00 [0.00, 4.00] | 0.00 [0.00, 2.50] | 0.006 |
| Predominant variant (%) | | | <0.001 |
| - omicron | 102 (6.8) | 17 (16.5) | |
| - omicron_BA.1 | 224 (14.9) | 29 (28.2) | |
| - omicron_BA.2 | 408 (27.1) | 16 (15.5) | |
| - omicron_BA.5 | 760 (50.5) | 40 (38.8) | |
| - omicron_BQ.1 | 5 (0.3) | 0 (0.0) | |
| - omicron_XBB.1.5 | 5 (0.3) | 1 (1.0) | |

*IQR: interquartile range; COPD: chronic obstructive pulmonary disease; CKD: chronic kidney disease*
